# Severe Infections as a Novel Risk Enhancing Factor for Incident Heart Failure

**DOI:** 10.1101/2025.11.12.25340120

**Authors:** Ryan T. Demmer, Brandon R. Grossardt, Sadiya S. Khan, Barry A. Borlaug

## Abstract

**Background:** Severe infections might be a novel risk marker for incident heart failure (HF). We examined the relationship between infection-related hospitalization (IRH) and HF in a large clinical health system and assessed whether adding IRH improved predictive utility of contemporary HF risk equations.

**Methods:** We studied 80,880 adults (14,150 with an IRH, 66,730 age- and sex-matched comparators) aged ≥18 years without HF at baseline in the Rochester Epidemiology Project (REP) between 1/1/2005 – 12/31/2019. IRH was identified using select International Classification of Disease (ICD) codes from hospital discharge records. HF incidence was defined as ≥1 HF-related ICD codes. We used multivariable-adjusted Cox proportional hazards models to assess the association between IRH and incident HF. In a separate cohort of 29,219 adults aged 30-79 years without baseline CVD in the REP, we calculated Harrell’s C-statistic to assess whether IRH information improved discrimination of the PREVENT-HF equations.

**Results:** Among 80,880 adults, 53.3% were women, 85.6% were White, 95.1% were non-Hispanic, and mean (SD) age at index was 56.0 (20.0) years. Median follow-up time was 6.2 years. Among the IRH group, 2,729 had an incident HF event (2.51 per 100 person years) versus 4,854 with an incident HF event (0.98 per 100 person years) in the comparator group. After multivariable adjustment, the hazard ratio (HR) for incident HF among participants who had an IRH compared to those who did not was 1.50 (95% CI: 1.43, 1.58). This relationship was consistent across different types of infections. In a separate cohort, the addition of IRH to PREVENT-HF increased the C-statistic from 0.776 to 0.779 (change 0.003 [0.001-0.004]). The PREVENT-HF risk score underestimated the 10-year risk of HF by up to 9% among participants with an IRH.

**Conclusion:** IRH was independently associated with incident HF but provided negligible change to overall 10-year HF risk discrimination. However, among individuals with an IRH, incorporating this information meaningfully improved 10-year HF risk prediction, suggesting that severe infections may serve as a risk-enhancing factor warranting enhanced preventive measures, particularly in populations where IRH is more common.

**Clinical Perspective:** *What’s New?:*

- We found that infection-related hospitalizations, obtained through electronic health records, were associated with incident heart failure.
- In a large clinical system in the upper Midwest, the PREVENT-HF equations, calculated from data available in electronic health records (EHR), accurately predicted 10-year HF risk overall, but may modestly under-estimate risk among individuals with a prior infection related hospitalization.
- Incorporating EHR-derived information on a broad range of previous infection-related hospitalizations may support enhanced preventive measures as a risk-enhancing factor.

*Clinical Implications:*

- Infection-related hospitalization (IRH) identifies adults at higher risk for developing heart failure, suggesting that severe infection events could be viewed as risk-ehnacing factors that aid in indentifying patients who may need closer follow-up and prevention efforts.
- Incorporating IRH into sequential risk assessment frameworks beginning with the PREVENT-HF equations highlights personalized opportunities for prevention for patients recently hospitalized with infection.

---

Heart failure (HF) is a complex clinical syndrome with significant impact on healthcare costs and population health. Estimates suggest that by 2030 more than 8 million people will be living with HF in the United States, and the medical costs of HF will double in the next several decades.^1^

Recent results from clinical trials demonstrate encouraging benefits of new medications, including sodium glucose co-transporter-2 inhibitors (SGLT2i) and glucagon like peptide-1 receptor agonists (GLP1-RA), for patients with prevalent HF.^2–5^ These same medicines also reduce the risk for new-onset HF in large outcome trials that included patients without prevalent HF at the time of enrollment.^6–8^ In light of these advances, there is a need to better identify population subgroups who may benefit most from the use of these medications in order to prioritize treatment given cost and access considerations. Current risk prediction models exist to guide clinical decision making.^9^ However, the major risk factors for HF account for only 52% of incident HF events^1^ suggesting that additional novel risk factors may enhance risk prediction and identify high risk groups potentially eligible for emerging HF prevention therapies.

Severe infections have long been linked to increased risk for coronary artery disease and stroke. Recent evidence suggests that severe infections also increase HF risk.^10, 11^ However, there are no studies using information on infection history obtained from electronic health records limiting knowledge about the clinical and predictive utility of this information. Such information would directly inform the relevance of considering infection history as a risk-enhancing factor^12^ to augment risk stratification and clinical decision making. Moreover, if knowledge about history of severe infections enhances risk prediction, new research opportunities may exist to study whether the enhanced risk may be mitigated with more aggressive and/or earlier treatment of traditional risk factors after infection.

In this article, we first evaluate the association between severe infections and risk of incident HF in the geographically-defined population of Olmsted County, Minnesota using the resources of the Rochester Epidemiology Project (REP), a records-linkage system that allows for population-based risk estimation. Second, we evaluated whether incorporating information on recent severe infections improved the predictive performance of the 10-year American Heart Association PREVENT^TM^-HF equations.

## METHODS

### Study Population

The Rochester Epidemiology Project (REP) is a medical records linkage system that enables population-based research in Olmsted County, Minnesota and the surrounding areas.^13, 14^ The REP is unique through its use of healthcare visits to construct a residency census, which allows for the complete enumeration of geographically-defined populations.^15^ Our analysis had two goals: 1) test the etiological hypothesis that severe infections are associated with risk for incident HF development in a REP subcohort (called etiologic cohort); and 2) test whether adding information about a recent severe infection to the PREVENT-HF equations improved the predictive performance in a second REP subcohort (called clinical risk prediction cohort). Formation of the two distinct cohorts is described below.

### Etiologic Cohort

For the purpose of assessing the association between severe infections and incident HF within the REP, we designed a population-based historical cohort comprised of individuals with an infection-related hospitalization (IRH; the exposure) and a comparator group of age- and sex-matched individuals from the same geographically-defined population without a recent IRH in the five years before the date of the index infection. We refer to this as the ‘etiologic cohort’. Individuals with any ICD diagnosis codes for HF at any time before the index date (i.e., prevalent HF) were excluded from the cohort. The IRH group was identified using infection-related hospitalization discharge diagnosis ICD codes (**Supplemental Table 1**) in the primary or in the first five secondary diagnosis positions and the following inclusion criteria: i) the first occurrence of a hospitalization occurring from 1/1/2005 to 12/31/2019; ii) resident of Olmsted County, Minnesota on the date of hospital discharge; iii) age 18 years or older at the time of hospitalization discharge; iv) had no other infection-related hospitalizations in the 5 years before the anchoring index hospitalization (i.e., *de novo* infection, to exclude persons with frailty and/or unique underlying susceptibility to frequent recurrent infections); v) provided Minnesota Research Authorization permissions; and vi) had at least one health care visit after their IRH index date and on or before 12/31/2023 (end of study). For each person in the exposed infection-related hospitalization group, we identified up to 5 ***comparator persons*** using the following criteria to ensure that comparators: i) were of the same sex, similar age (± 1 year); ii) were residents of Olmsted County, Minnesota on the hospitalization discharge date of the exposed person (i.e., index date); iii) had at least one health care visit after their matched index date and on or before 12/31/2023 (end of study). Comparator persons were excluded if they had any HF history or any infection-related hospitalization in the 5 years before the matched index date. The Etiologic Cohort included 80,880 participants (14,150 with IRH and 66,730 comparators without IRH) and inclusion is described in **Figure 1, left panel**. HF outcomes were defined at the earliest date of HF diagnosis after index date using ICD-9 codes 428* and ICD-10 codes I50*. Persons with no clinical HF diagnoses after the index date were censored in Cox models at the earliest of (1) the most recent clinical visit date if before 12/31/2023 (lost to follow-up before end of study); (2) death; or (3) 12/31/2023 (end of study). Persons who died during follow-up were only included as a HF outcome when a clinical diagnosis of HF was given before death (i.e., we did not include HF mentioned only on death certificates).

**Figure 1.**
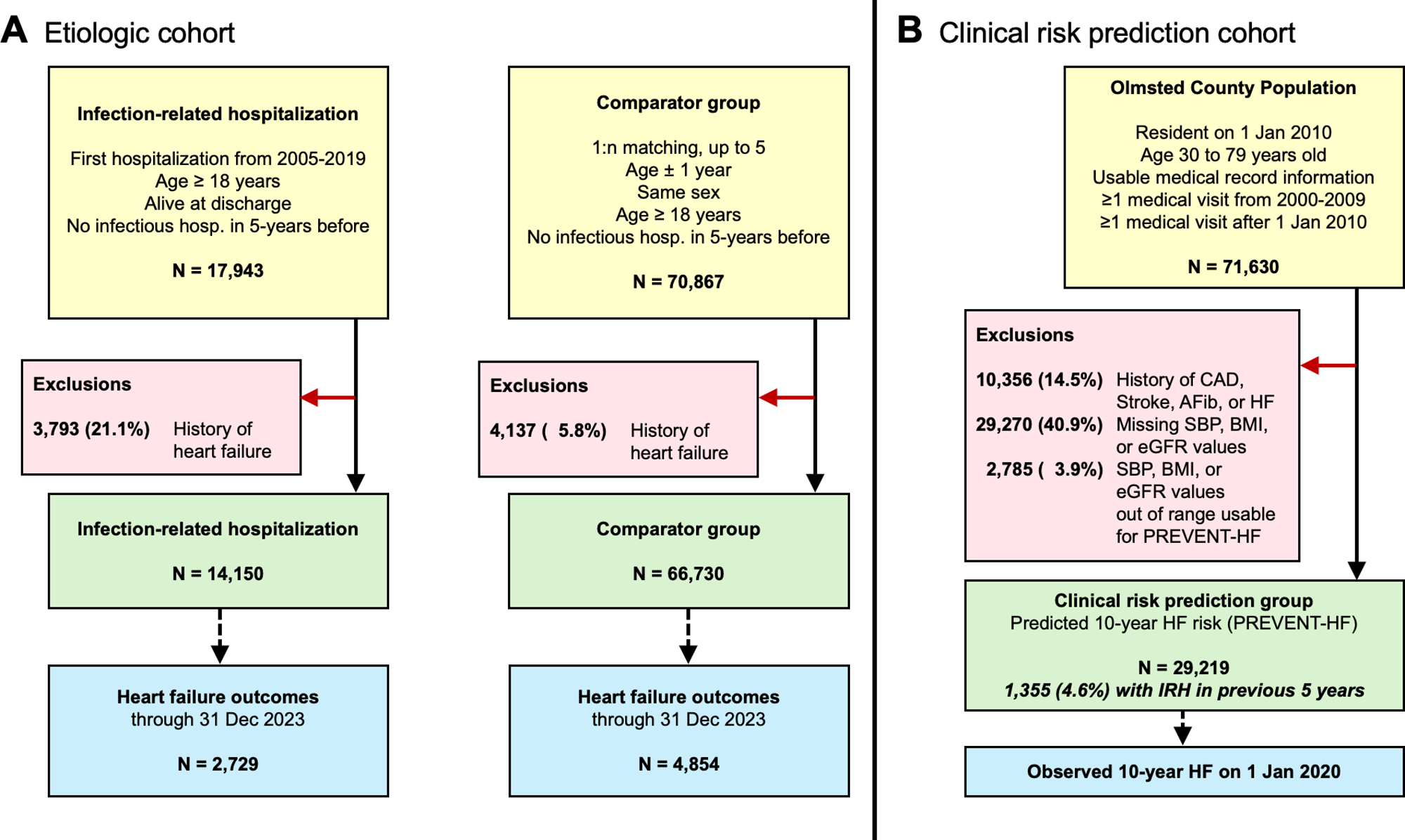
Participant flow chart for etiologic analyses (A) and clinical risk prediction (B).

### Clinical Risk Prediction Cohort

We additionally constructed a separate population-based ‘risk prediction’ cohort to determine whether recent IRH adds predictive utility to the 10-year PREVENT-HF equations. We identified 71,630 persons in the geographically-defined population of Olmsted County, MN meeting the following inclusion criteria: i) 30 to 79 years of age on 1/1/2010 (fixed baseline population); ii) provided Minnesota Research Authorization permissions; iii) had at least one health care visit for which PREVENT-HF risk score information was available in the 10 years before January 1^st^, 2010; iv) had at least one health care visit for which diagnostic information was available after January 1^st^, 2010 (i.e., at least some data for which HF outcome could be assessed). We excluded persons with: i) any ICD diagnosis code for coronary artery disease, stroke, atrial fibrillation, or HF before 1/1/2010; ii) missing risk factor values of systolic blood pressure, body mass index (BMI), or estimated glomerular filtration rate (eGFR) within the 3-year look back period from 1/1/2007 through 12/31/2009; iii) risk factor values that were outside of the range of those included in the creation of the PREVENT-HF equations (e.g., BMI ≥ 40 kg/m^2^) yielding 29,219 participants for analysis. Our inclusion/exclusion criteria were designed to match, as closely as possible, those used in the PREVENT equations development. A flow chart of inclusion is shown in **Figure 1, right panel and Supplemental Figure 1**.

Information used to calculate the 10-year PREVENT-HF risk was extracted from the electronic medical records. In particular, current smoking status was self-reported at the time of clinical visits. Diabetes status was assessed via ICD diagnosis codes, and use of anti-hypertensive medications was assessed via written prescription orders (a surrogate for actual use of medications). For continuous risk factors (systolic blood pressure, BMI, and eGFR), we used the value nearest to, but within the three years before the fixed inclusion date of 1 Jan 2010. Exclusions due to missing data are summarized in **Supplemental Figures 1 & 2.** Recent infection related hospitalization (IRH; our novel risk factor) was defined using the same hospital discharge diagnoses as in the Etiologic Cohort and included hospitalizations for the 5-year period from 1 Jan 2005 to 31 Dec 2009 (i.e., the 5 years before the fixed inclusion date of 1 Jan 2010).

## Statistical analyses

All statistical analyses were performed using SAS v. 9.4M8 and statistical significance was defined using the conventional two-tailed alpha level of 0.05. Summary statistics of baseline characteristics are reported as mean (SD) or median (interquartile intervals) and frequencies as appropriate.

### Etiologic Cohort: Risk of HF after severe infection

HF risk was assessed using Cox time-to-event regression models with age as the time scale. We present both unadjusted Cox model results and results adjusted for sex (male vs. female), race (White vs. non-White), ethnicity (non-Hispanic vs. Hispanic), education (Less than 4-year college education vs. 4-year college education or higher vs. unknown education), quinquennia of calendar year at index (calendar years 2005-2009 and calendar years 2010-2014; reference = 2015-2019), baseline BMI (underweight, overweight, obesity class I, obesity class II, obesity class III, unknown BMI vs. normal weight), baseline smoking status (former smoker, current smoker vs. never smoker), and baseline prevalent hypertension, diabetes, coronary artery disease, or atrial fibrillation. We tested for evidence of effect modification by sex and age group using Cochrane’s Q-test for heterogeneity of hazards ratios.

### Clinical Risk Prediction Cohort: Utility of IRH for 10-year HF risk prediction

We assessed the calibration and discrimination of the 10-year PREVENT-HF equations in the separate Clinical Risk Prediction Cohort. We compared the predicted 10-year HF risk to the observed 10-year HF risk in 10 strata defined by decile of predicted risk (i.e., predicted vs. observed 10-year risk of HF). To assess the added predictive value of incorporating recent IRH to the 10-year PREVENT-HF risk, we regressed the log-odds of incident HF on the PREVENT-HF log-odds score plus an indicator variable for presence of an IRH in the 5-years before 1 Jan 2010 to determine whether the IRH-informed model demonstrated stronger agreement with the observed risk when compared to the original PREVENT-HF predicted risk. Change in model discrimination with the addition of knowledge about recent IRH was assessed using change in Harrell C-statistic. Follow-up time was computed as time between 1 Jan 2010 and either incident HF, last contact with the medical system (lost to follow-up), death, or complete follow-up through 1/1/2020 free of HF.

## RESULTS

### Etiologic Cohort

Characteristics of persons included in the etiologic cohort are presented in **Table 1**. Participants were a mean age at index of 56.0 years (SD=20.0), 85.6% white, 95.1% nonHispanic, and 53.3% female. Those with IRH were older (mean age = 57.8 vs. 55.6) and more likely to be female (54.4% vs. 53.1%). Half of all hospitalized infections were classified as either respiratory (28.8%) or urinary tract infections (22.3%). Among 606,202 person years of follow-up, the incidence density of HF was 12.5 per 1,000 person years. The cumulative incidence of HF was 7,583/80,880 (9.4%) during a median follow-up of 6.2 years (interquartile interval = 3.1 to 11.1 years). Among those with IRH, the median (interquartile interval) follow-up time between IRH and incident HF was 3.5 (1.1 to 7.1) years.

**Table 1.** Characteristics of persons in the infection-related hospitalization and comparator groups.

| Characteristic | Infection-related hospitalization (IRH) group |  | Comparator group |  | P-value difference |
| --- | --- | --- | --- | --- | --- |
|  | N | % | N | % |  |
| **All included persons** | 14,150 | --- | 66,730 | --- | --- |
| **Sex** |  |  |  |  | 0.005 |
| Female | 7,700 | 54.4 | 35,453 | 53.1 |  |
| Male | 6,450 | 45.6 | 31,277 | 46.9 |  |
| **Age at index, years \*** |  |  |  |  | <.0001 |
| 18-39 | 3,480 | 24.6 | 17,094 | 25.6 |  |
| 40-59 | 3,898 | 27.5 | 20,150 | 30.2 |  |
| 60-79 | 4,170 | 29.5 | 21,311 | 31.9 |  |
| ≥ 80 | 2,602 | 18.4 | 8,175 | 12.3 |  |
| **Race** |  |  |  |  | <.0001 |
| White | 12,106 | 85.6 | 57,132 | 85.6 |  |
| Black | 834 | 5.9 | 2,800 | 4.2 |  |
| Asian | 518 | 3.7 | 3,100 | 4.6 |  |
| Other/mixed | 662 | 4.7 | 2,659 | 4.0 |  |
| Unknown | 30 | 0.2 | 1,039 | 1.6 |  |
| **Hispanic ethnicity** |  |  |  |  | 0.49 |
| Non-Hispanic | 13,467 | 95.2 | 63,416 | 95.0 |  |
| Hispanic | 683 | 4.8 | 3,314 | 5.0 |  |
| **Education level** |  |  |  |  | <.0001 |
| < 4 year college degree | 8,929 | 63.1 | 31,486 | 47.2 |  |
| ≥ 4 year college degree | 4,260 | 30.1 | 25,656 | 38.4 |  |
| Unknown | 961 | 6.8 | 9,588 | 14.4 |  |
| **Calendar year at index** |  |  |  |  | <.0001 |
| 2005-2009 | 4,691 | 33.2 | 20,795 | 31.2 |  |
| 2010-2014 | 4,495 | 31.8 | 20,694 | 31.0 |  |
| 2015-2019 | 4,964 | 35.1 | 25,241 | 37.8 |  |
| **BMI at index, kg/m<sup>2</sup>** |  |  |  |  | <.0001 |
| < 18.5 (Underweight) | 469 | 3.3 | 975 | 1.5 |  |
| 18.5 to <25 (Normal weight) | 4,196 | 29.7 | 19,234 | 28.8 |  |
| 25 to <30 (Overweight) | 4,364 | 30.8 | 22,336 | 33.5 |  |
| 30 to <35 (Class I Obesity) | 2,650 | 18.7 | 12,983 | 19.5 |  |
| 35 to <40 (Class II Obesity) | 1,315 | 9.3 | 5,322 | 8.0 |  |
| 40+ (Class III Obesity) | 1,099 | 7.8 | 3,272 | 4.9 |  |
| Unknown | 57 | 0.4 | 2,608 | 3.9 |  |
| **Smoking status at index** |  |  |  |  | <.0001 |
| Never smoker | 5,555 | 39.3 | 37,834 | 56.7 |  |
| Former smoker | 5,768 | 40.8 | 21,689 | 32.5 |  |
| Current smoker | 2,827 | 20.0 | 7,207 | 10.8 |  |
| **Other conditions at index** |  |  |  |  |  |
| Hypertension | 5,745 | 40.6 | 18,594 | 27.9 | <.0001 |
| Diabetes | 3,504 | 24.8 | 11,150 | 16.7 | <.0001 |
| Coronary artery disease | 1,603 | 11.3 | 4,476 | 6.7 | <.0001 |
| Atrial fibrillation | 715 | 5.1 | 1,784 | 2.7 | <.0001 |
| **Type of infection** |  |  |  |  | --- |
| All infections of any type | 14,150 | 100.0 | --- | --- |  |
| Respiratory | 4,080 | 28.8 | --- | --- |  |
| Influenza subset | 312 | 2.2 | --- | --- |  |
| Urinary tract (UTI) | 3,160 | 22.3 | --- | --- |  |
| Digestive tract | 2,079 | 14.7 | --- | --- |  |
| Skin | 1,921 | 13.6 | --- | --- |  |
| Blood / circulatory system | 1,167 | 8.2 | --- | --- |  |
| Hospital-acquired | 465 | 3.3 | --- | --- |  |
| All other infections | 5,141 | 36.3 | --- | --- |  |

\* The index ages of included persons as mean (standard deviation) were 57.8 (21.1) for the IRH group and 55.6 (19.7) for the comparator group (p < 0.0001).

### Association between infection and incident HF

IRH was associated with increased risk of incident HF (**Table 2 & Figure 2**). The HR for incident HF among those with vs. without IRH was 1.50 (95% CI: 1.43, 1.58) after multivariable adjustment. This relationship was strongest among individuals 18-39 years of age at index, and the strength of relative hazards decreased monotonically with older age, but remained significant in all age groups except among those aged ≥80 years.

**Figure 2.**
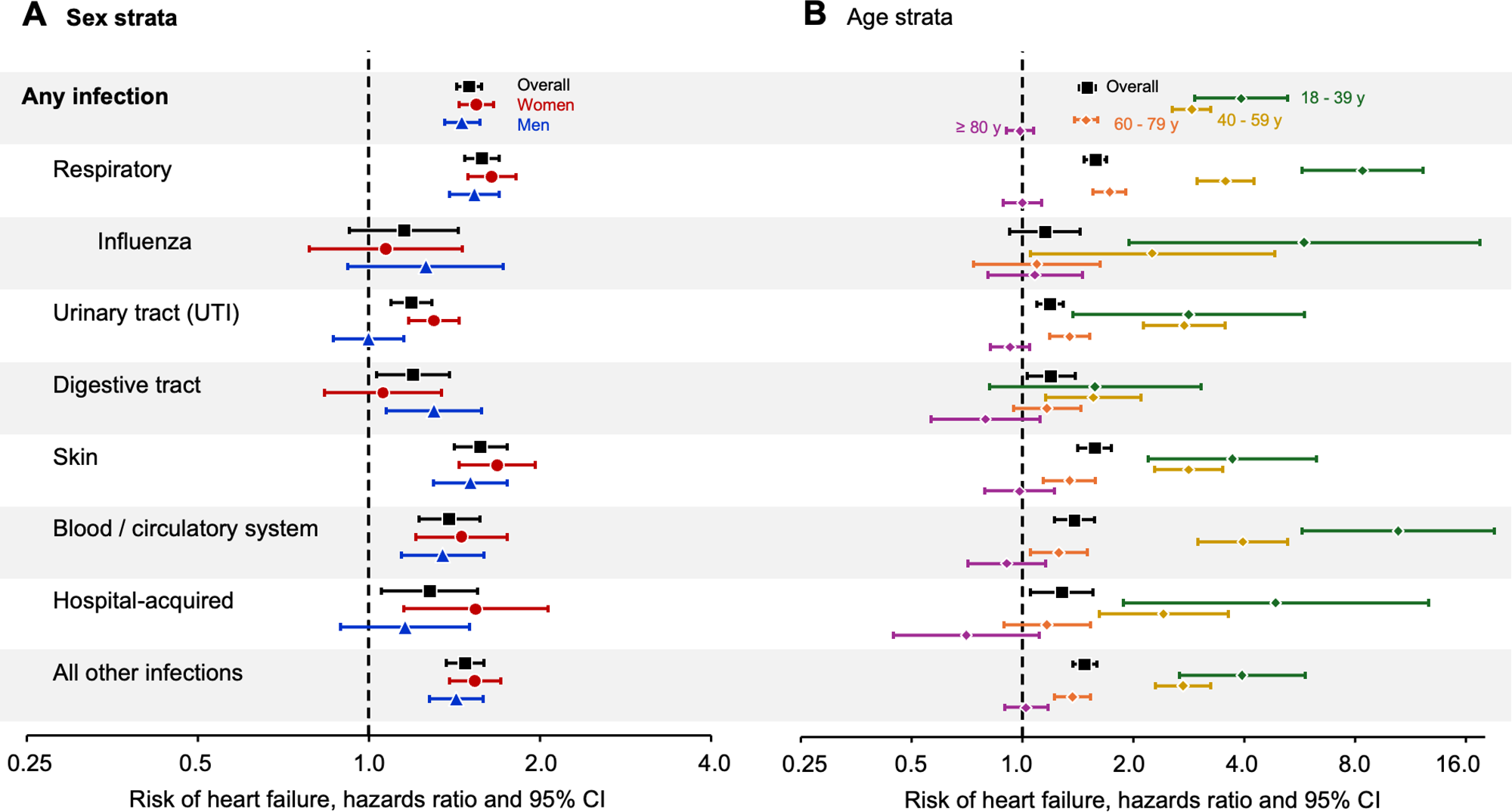
Relative risk (hazard ratios) of heart failure after infection-related hospitalization, separate by infection type category and stratified by sex (A) and age (B).

**Table 2.** Risk of heart failure after infection-related hospitalization.

| Description | Persons | Person years | Heart failure N (incid.*) | Risk of heart failure |  |  |  |
| --- | --- | --- | --- | --- | --- | --- | --- |
|  |  |  |  | Unadjusted model † |  | Adjusted model ‡ |  |
|  |  |  |  | HR (95% CI) | p-value | HR (95% CI) | p-value |
| **Overall pooled analyses** |  |  |  |  |  |  |  |
| IRH group | 14,150 | 108,755 | 2,729 (2.51) | **1.81 (1.73 to 1.90)** | **<.0001** | **1.50 (1.43 to 1.58)** | **<.0001** |
| Comparator group | 66,730 | 497,447 | 4,854 (0.98) | 1.0 (referent) | --- | 1.0 (referent) | --- |
| **Age at index strata** |  |  |  |  |  |  |  |
| **18-39 y** |  |  |  |  |  |  |  |
| IRH group | 3,480 | 32,094 | 112 (0.35) | **5.92 (4.48 to 7.83)** | **<.0001** | **3.93 (2.94 to 5.26)** | **<.0001** |
| Comparator group | 17,094 | 144,166 | 88 (0.06) | 1.0 (referent) | --- | 1.0 (referent) | --- |
| **40-59 y** |  |  |  |  |  |  |  |
| IRH group | 3,898 | 36,085 | 523 (1.45) | **3.94 (3.51 to 4.41)** | **<.0001** | **2.88 (2.56 to 3.25)** | **<.0001** |
| Comparator group | 20,150 | 189,166 | 698 (0.37) | 1.0 (referent) | --- | 1.0 (referent) | --- |
| **60-79 y** |  |  |  |  |  |  |  |
| IRH group | 4,170 | 29,966 | 1,215 (4.05) | **1.83 (1.70 to 1.96)** | **<.0001** | **1.49 (1.39 to 1.61)** | **<.0001** |
| Comparator group | 21,311 | 138,847 | 2,419 (1.74) | 1.0 (referent) | --- | 1.0 (referent) | --- |
| **≥ 80 y** |  |  |  |  |  |  |  |
| IRH group | 2,602 | 10,610 | 879 (8.28) | **1.12 (1.03 to 1.21)** | **0.008** | 0.99 (0.91 to 1.08) | 0.79 |
| Comparator group | 8,175 | 25,269 | 1,649 (6.53) | 1.0 (referent) | --- | 1.0 (referent) | --- |
| **Women only** |  |  |  |  |  |  |  |
| IRH group | 7,700 | 60,320 | 1,460 (2.42) | **1.88 (1.76 to 2.01)** | **<.0001** | **1.55 (1.44 to 1.66)** | **<.0001** |
| Comparator group | 35,453 | 267,005 | 2,341 (0.88) | 1.0 (referent) | --- | 1.0 (referent) | --- |
| **Age at index strata** |  |  |  |  |  |  |  |
| **18-39 y** |  |  |  |  |  |  |  |
| IRH group | 1,955 | 18,321 | 59 (0.32) | **8.01 (5.27 to 12.18)** | **<.0001** | **4.90 (3.13 to 7.65)** | **<.0001** |
| Comparator group | 9,558 | 81,190 | 35 (0.04) | 1.0 (referent) | --- | 1.0 (referent) | --- |
| **40-59 y** |  |  |  |  |  |  |  |
| IRH group | 2,000 | 19,158 | 262 (1.37) | **5.38 (4.52 to 6.41)** | **<.0001** | **3.48 (2.89 to 4.19)** | **<.0001** |
| Comparator group | 10,065 | 97,412 | 247 (0.25) | 1.0 (referent) | --- | 1.0 (referent) | --- |
| **60-79 y** |  |  |  |  |  |  |  |
| IRH group | 2,129 | 16,058 | 613 (3.82) | **2.00 (1.81 to 2.21)** | **<.0001** | **1.61 (1.45 to 1.79)** | **<.0001** |
| Comparator group | 10,721 | 72,605 | 1,087 (1.50) | 1.0 (referent) | --- | 1.0 (referent) | --- |
| **≥ 80 y** |  |  |  |  |  |  |  |
| IRH group | 1,616 | 6,782 | 526 (7.76) | **1.12 (1.00 to 1.24)** | **0.046** | 1.01 (0.90 to 1.12) | 0.91 |
| Comparator group | 5,109 | 15,799 | 972 (6.15) | 1.0 (referent) | --- | 1.0 (referent) | --- |
| **Men only** |  |  |  |  |  |  |  |
| IRH group | 6,450 | 48,436 | 1,269 (2.62) | **1.74 (1.62 to 1.86)** | **<.0001** | **1.46 (1.36 to 1.57)** | **<.0001** |
| Comparator group | 31,277 | 230,442 | 2,513 (1.09) | 1.0 (referent) | --- | 1.0 (referent) | --- |
|  |  |  |  | Unadjusted model † |  | Adjusted model ‡ |  |
|  |  |  |  | HR (95% CI) | p-value | HR (95% CI) | p-value |
| **Age at index strata** |  |  |  |  |  |  |  |
| **18-39 y** |  |  |  |  |  |  |  |
| IRH group | 1,525 | 13,773 | 53 (0.38) | **4.58 (3.13 to 6.70)** | **<.0001** | **3.44 (2.31 to 5.12)** | **<.0001** |
| Comparator group | 7,536 | 62,976 | 53 (0.08) | 1.0 (referent) | --- | 1.0 (referent) | --- |
| **40-59 y** |  |  |  |  |  |  |  |
| IRH group | 1,898 | 16,927 | 261 (1.54) | **3.15 (2.71 to 3.67)** | **<.0001** | **2.50 (2.13 to 2.94)** | **<.0001** |
| Comparator group | 10,085 | 91,754 | 451 (0.49) | 1.0 (referent) | --- | 1.0 (referent) | --- |
| **60-79 y** |  |  |  |  |  |  |  |
| IRH group | 2,041 | 13,908 | 602 (4.33) | **1.68 (1.53 to 1.85)** | **<.0001** | **1.40 (1.26 to 1.55)** | **<.0001** |
| Comparator group | 10,590 | 66,242 | 1,332 (2.01) | 1.0 (referent) | --- | 1.0 (referent) | --- |
| **≥ 80 y** |  |  |  |  |  |  |  |
| IRH group | 986 | 3,827 | 353 (9.22) | 1.12 (0.98 to 1.28) | 0.09 | 0.96 (0.84 to 1.10) | 0.53 |
| Comparator group | 3,066 | 9,469 | 677 (7.15) | 1.0 (referent) | --- | 1.0 (referent) | --- |

CI = Confidence interval; HR = Hazards ratio; IRH = Infection-related hospitalization
\* Raw incidence of heart failure per 100 person years (number of heart failure outcomes divided by person years under observation).
† Unadjusted Cox time-to-event models use age as the time scale (i.e., start/stop interval coding), but do not adjust for other potential confounders.
‡ Adjusted Cox time-to-event models use age as the time scale and include 1/0 indicator adjustment variables for race (Black race, Asian race, Other/mixed race, and Unknown race), Hispanic ethnicity, education (4-year college education or higher and unknown education), calendar year at index quinquennia (calendar years 2005-2009 and calendar years 2010-2014), BMI at index (underweight, overweight, obesity class I, obesity class II, obesity class III, and unknown BMI), smoking status at index (former smoker and current smoker), and for the presence at index of the 4 conditions hypertension, diabetes, coronary artery disease, and atrial fibrillation. The overall pooled sex analyses also include male sex as an adjustment variable. Tests for heterogeneity of the HRs across age or sex strata were conducted using Cochrane's Q-test which yielded a p-value <0.0001 for age and a p-value of 0.2458 for sex.

This association was present and statistically significant across all types of infection classifications (**Table 3**). The relationship was modestly stronger for UTIs and hospital acquired infections among women (p for interaction <0.05, **Table 3**), whereas the association for digestive tract infections was stronger among men, but the test for effect modification was not statistically significant. Results overall and separately within type of IRH are shown graphically in **Figure 2**. Results for specific infection classifications within age and sex subgroups are presented in **Supplemental Tables 2A-2H**. Results investigating the absolute impact of IRH on the risk of HF are shown in **Figure 3** and demonstrate substantially greater absolute HF risk differences associated with infection, among older age groups.

**Figure 3.**
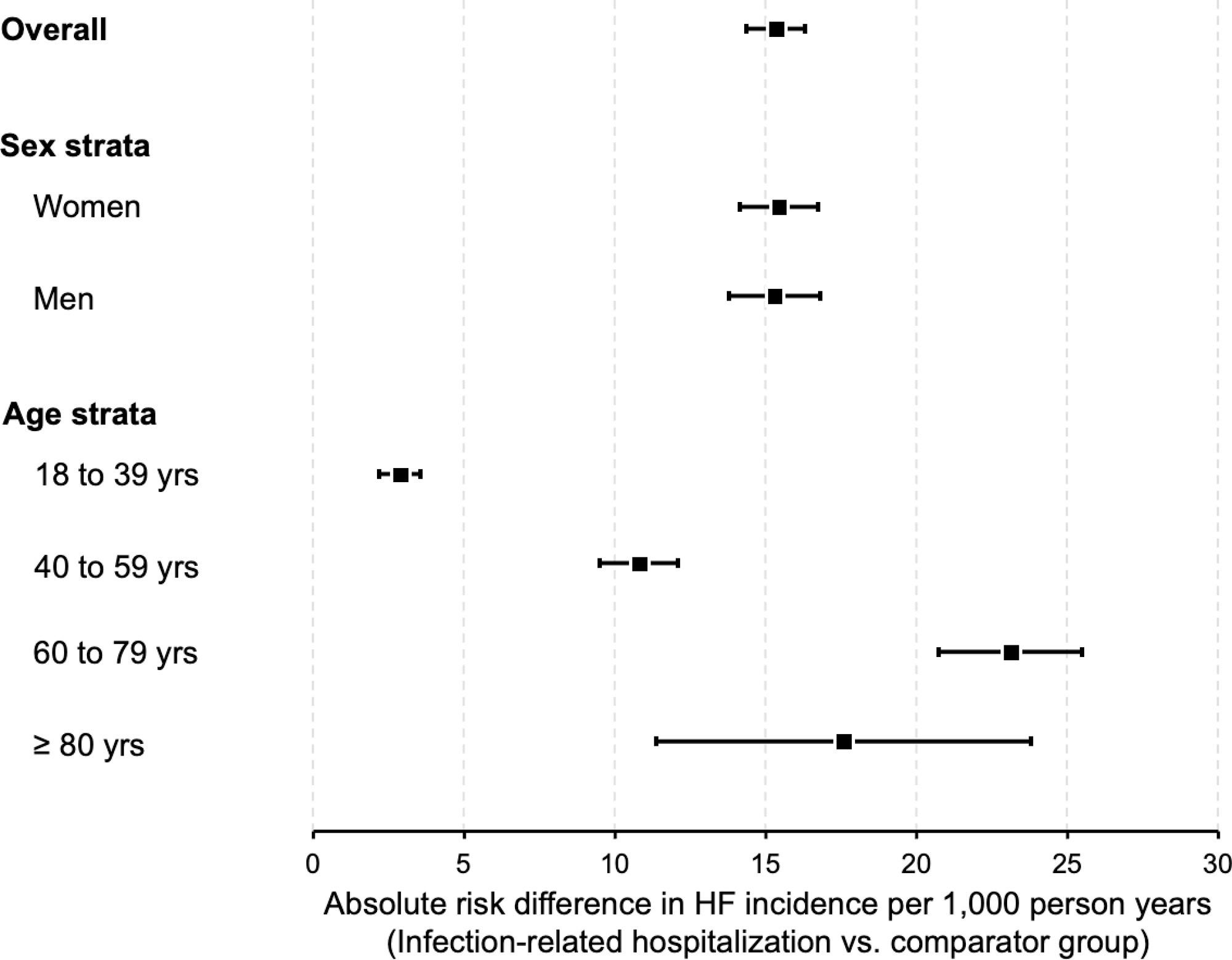
Absolute risk differences of heart failure after infection-related hospitalization, overall and stratified by sex and age.

**Table 3.** Risk of heart failure after infection-related hospitalization, separate by infection type category.

| Description | Persons | Person years | Heart failure N (incid.*) | Unadjusted model †<br>HR (95% CI) | p-value | Adjusted model ‡ §<br>HR (95% CI) | p-value |
| --- | --- | --- | --- | --- | --- | --- | --- |
| **Overall pooled analyses** |  |  |  |  |  |  |  |
| Comparator group | 66,730 | 497,447 | 4,854 (0.98) | 1.0 (referent) | --- | 1.0 (referent) | --- |
| **IRH group, type of infection** |  |  |  |  |  |  |  |
| Any infection | 14,150 | 108,755 | 2,729 (2.51) | 1.81 (1.73 to 1.90) | <.0001 | 1.50 (1.43 to 1.58) | <.0001 |
| Respiratory infection | 4,080 | 28,360 | 1,025 (3.61) | 1.94 (1.81 to 2.08) | <.0001 | 1.58 (1.48 to 1.70) | <.0001 |
| Urinary tract (UTI) | 3,160 | 21,491 | 698 (3.25) | 1.39 (1.28 to 1.51) | <.0001 | 1.19 (1.10 to 1.29) | <.0001 |
| Digestive tract | 2,079 | 19,674 | 181 (0.92) | 1.39 (1.20 to 1.62) | <.0001 | 1.20 (1.03 to 1.39) | 0.02 |
| Skin | 1,921 | 15,811 | 379 (2.40) | 2.19 (1.97 to 2.43) | <.0001 | 1.57 (1.42 to 1.75) | <.0001 |
| Blood / circulatory system | 1,167 | 8,772 | 278 (3.17) | 1.91 (1.69 to 2.15) | <.0001 | 1.39 (1.23 to 1.57) | <.0001 |
| Hospital-acquired | 465 | 4,158 | 107 (2.57) | 2.01 (1.66 to 2.44) | <.0001 | 1.28 (1.06 to 1.56) | 0.01 |
| All other infections | 5,141 | 38,445 | 838 (2.18) | 1.78 (1.65 to 1.92) | <.0001 | 1.48 (1.37 to 1.59) | <.0001 |
| **Women only §** |  |  |  |  |  |  |  |
| Comparator group | 35,453 | 267,005 | 2,341 (0.88) | 1.0 (referent) | --- | 1.0 (referent) | --- |
| **IRH group, type of infection** |  |  |  |  |  |  |  |
| Any infection | 7,700 | 60,320 | 1,460 (2.42) | 1.88 (1.76 to 2.01) | <.0001 | 1.55 (1.44 to 1.66) | <.0001 |
| Respiratory infection | 2,124 | 15,113 | 528 (3.49) | 2.01 (1.83 to 2.21) | <.0001 | 1.65 (1.49 to 1.82) | <.0001 |
| Urinary tract (UTI) | 2,280 | 16,014 | 480 (3.00) | 1.57 (1.42 to 1.74) | <.0001 | 1.30 (1.18 to 1.44) | <.0001 |
| Digestive tract | 1,001 | 9,695 | 72 (0.74) | 1.27 (1.01 to 1.61) | 0.045 | 1.06 (0.84 to 1.34) | 0.63 |
| Skin | 843 | 7,103 | 181 (2.55) | 2.23 (1.92 to 2.60) | <.0001 | 1.68 (1.44 to 1.97) | <.0001 |
| Blood / circulatory system | 561 | 4,469 | 123 (2.75) | 1.99 (1.66 to 2.39) | <.0001 | 1.46 (1.21 to 1.75) | <.0001 |
| Hospital-acquired | 215 | 1,873 | 47 (2.51) | 2.23 (1.67 to 2.97) | <.0001 | 1.54 (1.15 to 2.07) | 0.004 |
| All other infections | 2,776 | 21,503 | 440 (2.05) | 1.85 (1.67 to 2.05) | <.0001 | 1.54 (1.39 to 1.71) | <.0001 |
| **Men only §** |  |  |  |  |  |  |  |
| Comparator group | 31,277 | 230,442 | 2,513 (1.09) | 1.0 (referent) | --- | 1.0 (referent) | --- |
| **IRH group, type of infection** |  |  |  |  |  |  |  |
| Any infection | 6,450 | 48,436 | 1,269 (2.62) | 1.74 (1.62 to 1.86) | <.0001 | 1.46 (1.36 to 1.57) | <.0001 |
| Respiratory infection | 1,956 | 13,247 | 497 (3.75) | 1.83 (1.66 to 2.02) | <.0001 | 1.54 (1.39 to 1.70) | <.0001 |
| Urinary tract (UTI) | 880 | 5,477 | 218 (3.98) | 1.22 (1.06 to 1.41) | 0.005 | 1.00 (0.87 to 1.15) | 0.99 |
| Digestive tract | 1,078 | 9,979 | 109 (1.09) | 1.43 (1.18 to 1.74) | 0.0002 | 1.30 (1.08 to 1.58) | 0.007 |
| Skin | 1,078 | 8,708 | 198 (2.27) | 2.09 (1.80 to 2.41) | <.0001 | 1.51 (1.30 to 1.75) | <.0001 |
| Blood / circulatory system | 606 | 4,303 | 155 (3.60) | 1.74 (1.47 to 2.04) | <.0001 | 1.35 (1.14 to 1.60) | 0.0004 |
| Hospital-acquired | 250 | 2,285 | 60 (2.63) | 1.73 (1.34 to 2.23) | <.0001 | 1.16 (0.89 to 1.50) | 0.27 |
| All other infections | 2,365 | 16,942 | 398 (2.35) | 1.70 (1.53 to 1.89) | <.0001 | 1.43 (1.28 to 1.59) | <.0001 |

CI = Confidence interval; HR = Hazards ratio; IRH = Infection-related hospitalization
\* Raw incidence of heart failure per 100 person years (number of heart failure outcomes divided by person years under observation).
† Unadjusted Cox time-to-event models use age as the time scale (i.e., start/stop interval coding), but do not adjust for other potential confounders.
‡ Adjusted Cox time-to-event models use age as the time scale and include 1/0 indicator adjustment variables for race (Black race, Asian race, Other/mixed race, and Unknown race), Hispanic ethnicity, education (4-year college education or higher and unknown education), calendar year at index quinquennia (calendar years 2005-2009 and calendar years 2010-2014), BMI at index (underweight, overweight, obesity class I, obesity class II, obesity class III, and unknown BMI), smoking status at index (former smoker and current smoker), and for the presence at index of the 4 conditions hypertension, diabetes, coronary artery disease, and atrial fibrillation. The overall pooled sex analyses also include male sex as an adjustment variable.
§ Tests for differences in adjusted HRs between women and men were performed overall and in strata by infection type (p = 0.11 for any infection; p = 0.16 for respiratory infections; p < 0.0001 for urinary tract infections; p = 0.06 for digestive tract infections; p = 0.17 for skin infections; p = 0.40 for blood and circulatory system infections; p = 0.04 for hospital-acquired infections; and p = 0.16 for all other infections).

### Contribution of severe infection knowledge to clinic risk prediction

We included a total of 29,219 individuals from ages 30 to 79 years old on January 1^st^, 2010 in the clinical risk prediction cohort, 1,355 (4.6%) of whom had an IRH within 5 years priot to January 1^st^, 2010. Median (Q1, Q3) follow-up time for this analysis was 10 (10, 10) years, and the AUC for the 10-year risk of HF prediction using the PREVENT-HF equations were 0.776 (95%CI: 0.764 to 0.788) overall, 0.755 (0.737 to 0.773) among men, and 0.790 (0.773 to 0.807) among women. Overall, the PREVENT-HF equations were highly accurate and consistent with the observed risk of HF in both men and women (**Figure 4A**). However, when stratifying risk by history of recent IRH, the PREVENT-HF equations underestimated the observed 10-year risk by as much as 8.9% in men and 9.2% in women (**Figure 4B**). When incorporating recent history of IRH into the PREVENT-HF equations, the change in discrimination or AUC was 0.003 (0.001-0.004; p=0.0007 (**Figure 4C**). More details about the performance of the PREVENT-HF equations across strata by sex, age, and recent IRH can be found in **Supplemental Tables 3, 4, and 5**, respectively.

**Figure 4.**
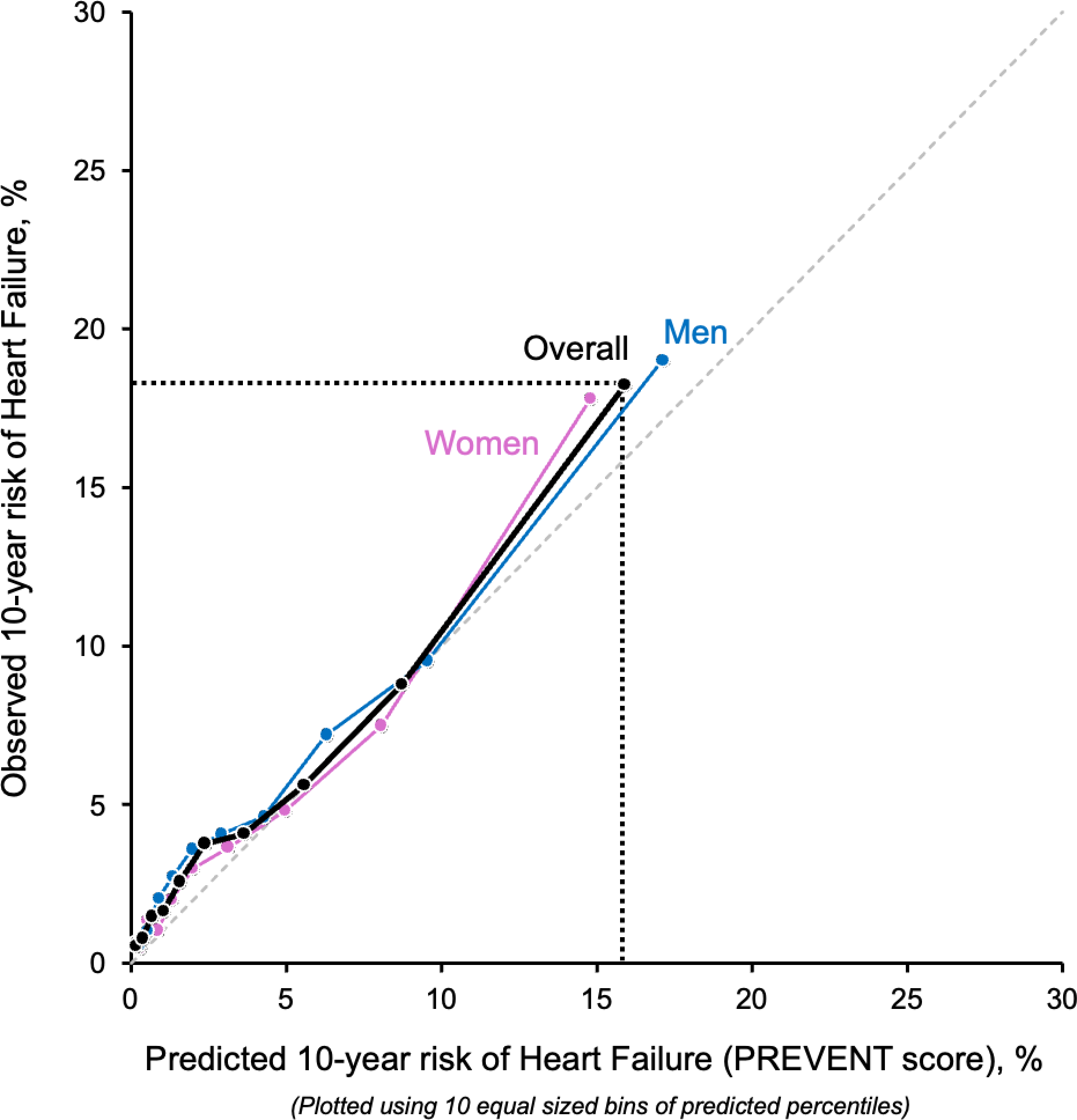
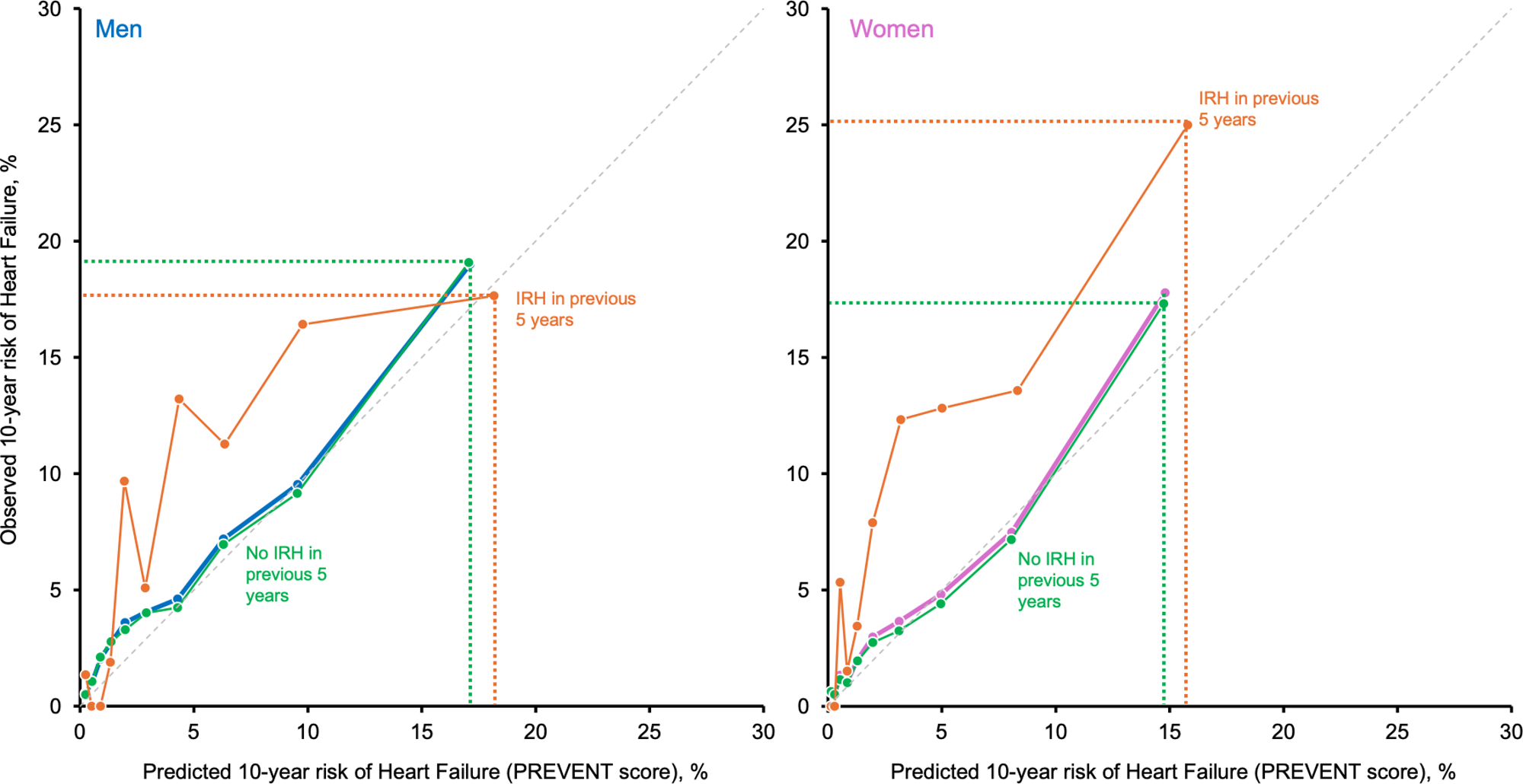
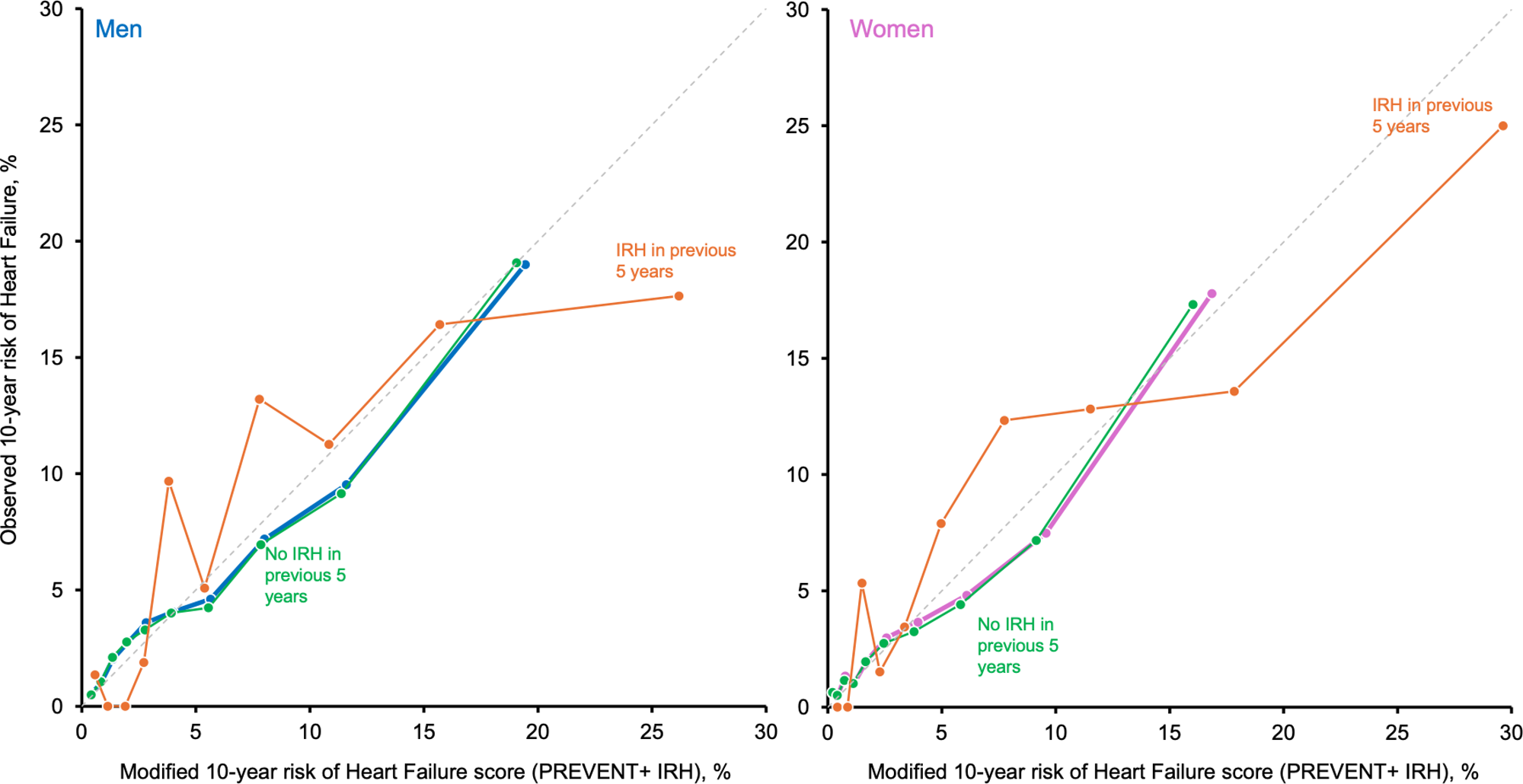
PREVENT score predicted versus observed 10-year risk of heart failure by decile of PREVENT score (A), among patients with vs. without infection related hospitalization (B), and among patients with vs. without severe infection after augmenting the PREVENT score with infection related hospitalization information (D). Results arise from 29,219 patients in the Rochester Epidemiology Project, 2010 - 2020.

## DISCUSSION

In a large midwestern population of adult men and women, we observed clinically meaningful associations between IRH and risk for incident HF during a median 6 years of longitudinal follow-up. Absolute risk differences were larger among older participants while relative differences were larger in younger participants. Results were consistent between men and women, with the exception of UTIs and blood/circulatory system infections, which were stronger in women, and digestive tract infections, which were stronger among men. In a separate clinical risk prediction cohort, the PREVENT-HF had good to excellent discrimination with minimal change in predictive utility as assessed with the lack of meaningful change in the C-statistic when IRH was added. However, among people with an IRH, the PREVENT-HF equations underestimated the observed HF risk by up to 9% suggesting potential clinical utility of IRH as a risk-enhancing factor.

To our knowledge, this is the first report to investigate whether the additional knowledge of IRH history is associated with incident HF and to evaluate the predictive utilty of IRH in the PREVENT-HF equations. This study used an innovative approach with an EHR-based system for identifying hospitalization events. Doing so in a real-world clinical health system increases the likelihood that these findings are clinically translatable and might have impact on preventive efforts.

The observed effect modification across age strata in which findings were strongest in the oldest age groups when using additive models (i.e., risk differences) but weakest in the oldest age groups when using multiplicative models (hazards ratios) is a common observation resulting from large differences in the absolute risk of HF across age strata. The large absolute risk differences among the old suggests that the greatest population impact of these findings is among older individuals because both the occurrence of severe infection and HF are high in this population.

### Biological plausibility and mechanisms

The initial response to severe infection is characterized by activation of the innate and adaptive immune systems, aimed at pathogen eradication. This involves the release of pro-inflammatory cytokines, activation of immune cells, and recruitment of these cells to the site of infection.^16^ Pathogen-associated molecular patterns (PAMPs) and damage-associated molecular patterns (DAMPs) released during infection can trigger pattern recognition receptors (PRRs) like Toll-like receptors (TLRs) on immune cells, initiating inflammatory cascades.^17^ Normally, this acute inflammation resolves upon pathogen clearance, restoring immune homeostasis. However, in severe infections, the inflammatory response can become dysregulated and non-resolving.

Several mechanisms promote the transition from acute to chronic immune activation after severe infection. First, persistent microbial antigens or non-resolving inflammation can chronically stimulate immune cells, leading to chronic cytokine production and immune cell hyperreactivity.^18^ Second, severe infections can induce immune exhaustion, characterized by dysfunctional immune cells with impaired effector functions but maintained or even increased expression of inhibitory receptors.^19^ Paradoxically, this state can co-exist with chronic inflammation, as exhausted immune cells may fail to effectively clear residual infection or dampen inflammatory signals, while still contributing to a state of dysregulated immunity.^20^ Furthermore, severe infections can disrupt the regulatory mechanisms of the immune system, such as regulatory T cells (T-regs), which are important for maintaining immune tolerance and resolving inflammation.^21^ Dysfunction or depletion of T-regs can contribute to uncontrolled inflammation and chronic immune activation post-infection.

Severe infections can also induce profound changes in DNA methylation patterns in various immune cell types resulting in epigenetic modifications. For example, studies have shown that sepsis can lead to widespread alterations in DNA methylation in monocytes, neutrophils, and T cells, affecting the expression of genes involved in inflammation, immune regulation, and cell metabolism.^22^ Other research has shown that sepsis can induce changes in histone acetylation at promoters of pro-inflammatory cytokine genes, contributing to prolonged inflammatory responses.^23^ Alterations in histone methylation patterns also have been linked to immune cell exhaustion and impaired antigen presentation capacity after severe infections.

Thinking beyond biological mechanisms directly linked to infection, it is possible that the hospitalization event itself (as a downstream consequence of infection) might explain the relationship between IRH and HF. Hospitalization is accompanied by a number of factors that might be related to waning health including immobility, isolation, loss of wages, and unexpected financial burdens or may serve as a proxy or surrogate for overall health status. Alternatively, discontinuation of medications for chronic conditions such as hypertension or hyperlipidemia might persist for months or years after hospitalization leading to increased risk of HF. Limited data exist formally exploring these explanations and future research is warranted in this area.

### Implications for HF risk stratification

Our findings may have implications for IRH as a potential risk-enhancing factor given the strong independent association between IRH and HF. However, the PREVENT-HF equations had good discrimination in this sample, which was consistent with other external validation samples. It is possible that the risk in this sample is likely underestimated due to our study design. Specifically, in the risk prediction cohort we only consider the role of infection within 5 years prior to baseline. Consequently, in our sample, the prevalence of recent IRH was only 4.6%, which is likely a substantial underestimation of the burden of severe infections during the adult life-course. For example, in the ARIC cohort, 47% of participants had an IRH during 30 years of follow-up.^10^ Data from the National Inpatient Survey show that among the top 20 principal diagnoses for inpatient stays, four are infection related with septicemia ranking first; collectively, these four infections represented 13.4% of inpatient stays in 2018.^24^ Therefore, the lifetime burden of severe infections is likely substantially greater than what is represented in our present analysis and the impact on risk prediction in clinical practice is likely also under-estimated.

### Strengths

Our study has some unique strengths. First, we use the Rochester Epidemiology Project (REP), a large real-world clinical population from a predominantly rural, midwestern health care system. Second, the REP captures essentially the entire Olmsted County population (∼160,000 individuals) and 98% of individuals in the REP have agreed to allow their health records to be used for health research.^14^ Accordingly, our results are generalizable to a large segment of the Midwest population. The REP includes data from multiple health systems, providing more complete information about the health status of individuals residing in our region. Other systems suffer from fragmentation of care because many people use multiple health systems for different purposes (primary care vs. emergency department visits vs. specialty care, etc.).

### Limitations

Our etiologic analyses are limited by a lack of detailed information on key confounders such as dietary behaviors, inflammatory biomarkers, and general measures of frailty which may predispose individuals to both severe infections and HF risk. It is possible that our results suffer from selection bias as we would not capture individuals in our comparator group who infrequently access the health care system. The study sample is largely white non-Hispanic; nevertheless, this sample is likely generalizable to a large segment of the U.S. Midwest. Moreover, the findings are supported by previous findings in large, diverse populations outside of the Midwest.

## Conclusions

We report that severe infections are associated with increased risk of HF in a large, population-based, healthcare system. In a separate sample, we show that knowledge of recent infection related hospitalization does not meaningfully improve discrimination but may hold promise to target early interventions to prevent HF as a key risk-enhancing factor. Future research is needed to better understand the underlying mechanisms connecting severe infection and HF, and how to operationalize strategies to prevent, diagnose, and treat the potential cardiac consequences of severe infections. Regardless of causality and mechanisms, there is an opportunity to enhance efforts in the primary prevention of HF in those with IRH.

## Funding Acknowledgments and Disclosures

Drs. Demmer and Borlaug receive research support from the National Institutes of Health (NIH). Dr. Borlaug receives research support from the United States Department of Defense, as well as research grant funding from AstraZeneca, Axon, Corvia, Novo Nordisk, and Tenax Therapeutics. Dr. Borlaug has served as a consultant for Actelion, Amgen, Aria, Axon Therapies, BD, Boehringer Ingelheim, Cytokinetics, Edwards Lifesciences, Lilly, Imbria, Janssen, Merck, Novo Nordisk, NGM, NXT, and VADovations, and is named inventor (US Patent no. 10,307,179) for the tools and approach for a minimally invasive pericardial modification procedure to treat heart failure. This work was also supported by the National Institute on Aging, AG 062580, and used the resources of the Rochester Epidemiology Project (REP) medical records-linkage system, which is supported by the National Institute on Aging, AG 058738, by the Mayo Clinic Research Committee, and by fees paid annually by REP users. The funding sources played no role in the design of the study, the analysis, the interpretation of study results, or the writing of the manuscript.

## Data Availability

Data can be made available upon request to the corresponding author and approval from the Mayo Clinic IRB and legal counsel. All requestors must be appropriately credentialed and have documented completed CITI training in human subjects research.

